# Hydroxychloroquine inhibits trained immunity – implications for COVID-19

**DOI:** 10.1101/2020.06.08.20122143

**Authors:** Nils Rother, Cansu Yanginlar, Rik G.H. Lindeboom, Siroon Bekkering, Mandy M.T. van Leent, Baranca Buijsers, Inge Jonkman, Mark de Graaf, Marijke Baltissen, Lieke A. Lamers, Niels P. Riksen, Zahi A. Fayad, Willem J.M. Mulder, Luuk B. Hilbrands, Leo A.B. Joosten, Mihai G. Netea, Michiel Vermeulen, Johan van der Vlag, Raphaël Duivenvoorden

**Affiliations:** Department of Nephrology, Radboud Institute for Molecular Life Sciences, Radboud university medical center, Nijmegen, The Netherlands; Department of Molecular Biology, Faculty of Science, Radboud Institute for Molecular Life Sciences, Oncode Institute, Radboud University Nijmegen, Nijmegen, the Netherlands; Department of Internal Medicine, Radboud Institute for Molecular Life Sciences, Radboud University Medical Center, Nijmegen, The Netherlands; Biomedical Engineering and Imaging Institute, Icahn School of Medicine at Mount Sinai, New York, NY, USA; Department of Medical Biochemistry, Amsterdam University Medical Centers, Amsterdam, The Netherlands; Laboratory of Chemical Biology, Department of Biomedical Engineering and Institute for Complex Molecular Systems, Eindhoven University of Technology, Eindhoven, The Netherlands; Department of Oncological Sciences, Icahn School of Medicine at Mount Sinai, New York, NY, USA; Department of Immunology and Metabolism, Life and Medical Sciences Institute (LIMES), University of Bonn, Bonn, Germany

## Abstract

SARS-CoV-2 infection can cause severe disease for which currently no specific therapy is available. The use of hydroxychloroquine to prevent or treat SARS-CoV-2 infection is controversial and its mode of action poorly understood. We demonstrate that hydroxychloroquine inhibits trained immunity at the functional and epigenetic level and is accompanied by profound changes in the cellular lipidome as well as reduced expression of interferon-stimulated genes. Trained immunity comprises a functional adaptation induced by epigenetic reprogramming which facilitates the anti-viral innate immune response. Our findings therefore suggest that hydroxychloroquine may not have a beneficial effect on the anti-viral immune response to SARS-CoV-2.

## INTRODUCTION

The severe acute respiratory syndrome coronavirus 2 (SARS-CoV-2), which causes coronavirus disease 2019 (COVID-19), has spread globally since the December 2019 outbreak in China. The majority of COVID-19 patients have mild symptoms, but some develop severe pneumonia^1^. The factors that cause severe illness are not fully understood, but a growing body of evidence points to an inadequate immune response and previous studies have shown that coronaviruses have multiple strategies to evade innate immune sensing^2^. This is exemplified by the fact that in COVID-19 patients a decreased type I interferon (IFN) response is observed, which is associated with impaired viral clearance^3,4^. Ineffective clearance of SARS-CoV-2 may lead to uncontrolled tissue inflammation and poor outcome.

To date, no specific therapy is available to treat COVID-19. The antimalarial drugs chloroquine and hydroxychloroquine have been proposed as therapeutic agents^5–7^. These drugs were observed to inhibit SARS-CoV-2 viral replication *in vitro* in primate cells^8^. However, there is no confirmation so far that these drugs can affect viral replication *in vivo* in humans^9^. Chloroquine and hydroxychloroquine also have immunomodulating properties, which may influence the disease course of COVID-19^10^. The use of chloroquine and hydroxychloroquine for the treatment of COVID-19 is controversial however, as there is no clear evidence of their efficacy and a poor understanding of their mode of action^11,12^. Better knowledge of how these 4-aminoquinolines affect the immune response is fundamentally important to uncover whether these drugs can, or cannot, be beneficial in the treatment of COVID-19.

In the current study, we investigated the immune response in COVID-19 and the immunomodulatory properties of hydroxychloroquine. Using an integrative approach with functional and transcriptomic analyses, we show marked alterations in function and phenotype of monocytes isolated from COVID-19 patients, and show interferon-stimulated genes to be associated with disease severity. By combining transcriptomic, metabolomic and epigenetic studies, we reveal that hydroxychloroquine can prevent the induction of trained immunity. Trained immunity is a functional adaptation of monocytes induced by epigenetic reprogramming that potentiates their anti-viral response^13^. Our findings suggest that hydroxychloroquine may not be favorable for the anti-viral immune response to SARS-CoV-2.

## RESULTS

### Monocyte phenotype and function in COVID-19

We studied 13 patients who were admitted to Radboud University Medical Center, a tertiary care university hospital, with a SARS-CoV-2 infection. Patients were included if they were older than 18 years of age and diagnosed with COVID-19. Blood was obtained at admission and five days after admission in patients who were still hospitalized. Treatment with chloroquine was started at time of admission and continued for five days. Median age was 68 years (IQR 54 – 73). Five patients had a history of pulmonary disease, three of cardiovascular disease, and three of malignancy. Most patients presented with fever (62%), cough (77%), and/or dyspnea (54%). Seven of the 13 patients required oxygen supplementation at presentation (all ≤ 5L/min). All patients had signs of pneumonitis on chest imaging. None of the patients were critically ill at the time of presentation. Patient characteristics are shown in **Extended Data Table 1**, and complete blood count of all subjects are shown in **Extended Data Table 2**.

We investigated the immune response in COVID-19 patients and compared it to healthy controls. For this purpose, peripheral blood mononuclear cells (PBMCs) were isolated from the blood and immune cell subsets were analyzed by flow cytometry (**Fig. 1a, Extended Data Fig. 1b,c**). At the time of admission, patients with COVID-19 had slightly fewer T lymphocytes, but no differences in other lymphocyte subsets (**Fig. 1b**). Monocytes were markedly increased in COVID-19 patients, mainly due to a striking increase in CD14^++^CD16^−^ (classical) monocytes (**Fig. 1c,d**). Interestingly, CD14^+^CD16^++^ (non-classical) monocytes were hardly detectable in COVID-19 patients (**Fig. 1d**). Human leucocyte antigen DR (HLA-DR) was reduced on monocytes from COVID-19 patients (**Fig. 1e**). Low HLA-DR expression was recently shown to be associated with monocyte hyperactivation and excessive release of interleukin-6 (IL-6) in COVID-19 patients^14^. Expression of CX3CR1, which is involved in monocyte chemotactic migration and is mostly expressed by the non-classical monocyte subset^15^, was reduced (**Fig. 1f**), in accordance with the observed decrease in non-classical monocytes (**Fig. 1d**). The integrin CD11b, a marker of monocyte activation, was upregulated on monocytes of COVID-19 patients (**Fig. 1g**). Lymphocyte and monocyte subsets as well as HLA-DR, CX3CR1 and CD11b expression did not change over the course of five days in patients who remained hospitalized (**Extended Data Fig. 2a-g**).

**Fig. 1.**
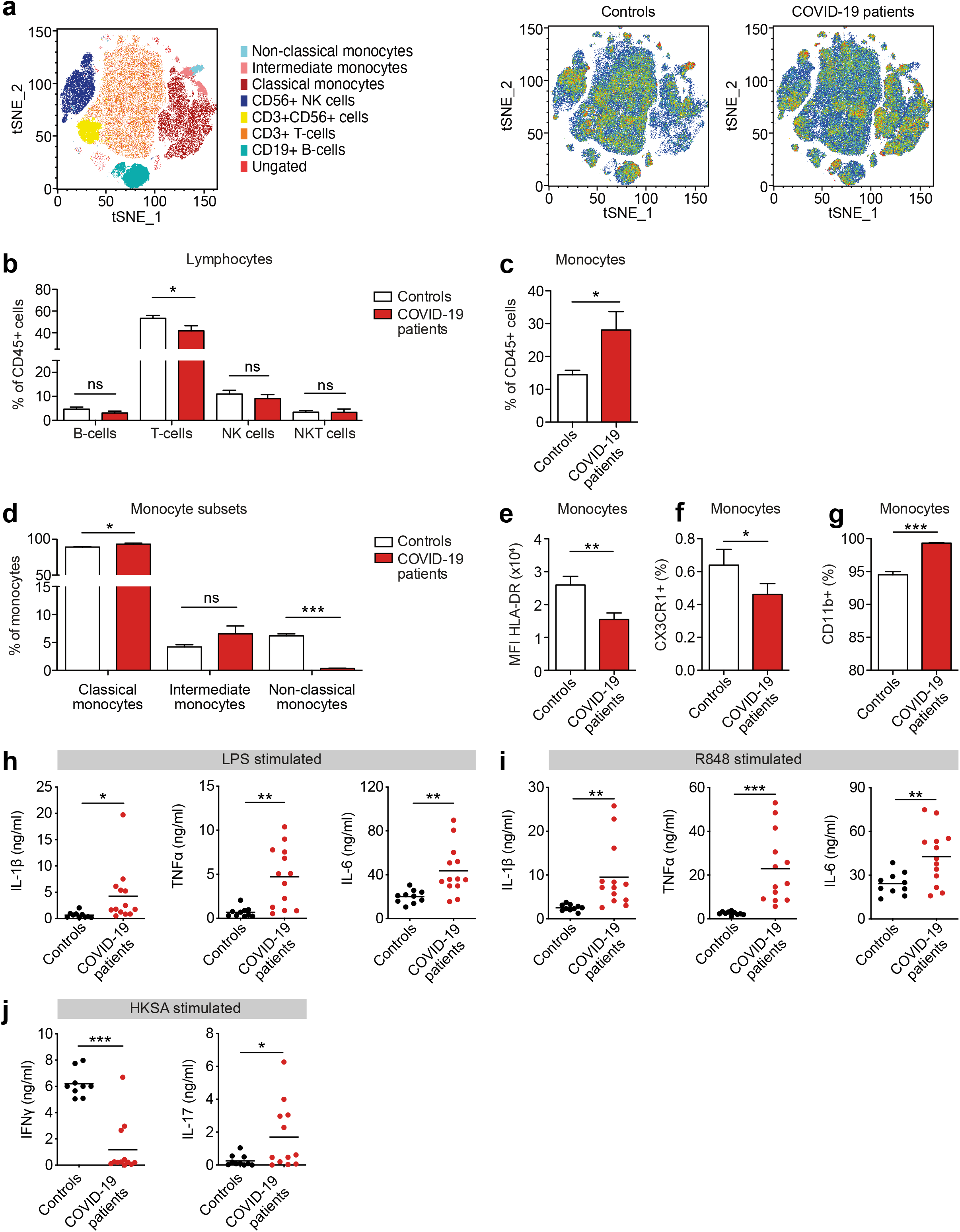
Innate immune response in COVID-19 patients at the time of admission. a-g, PBMCs isolated from COVID-19 patients at admission and from healthy controls were analyzed using flow cytometry (n = 10 for COVID-19 patients, n = 7 for healthy controls). a, tSNE plots showing unsupervised clustering on the expression of 10 markers (CD45, CD14, CD16, CD3, CD19, CD56, HLA-DR, CD11b, CCR2 and CX3CR1) in controls and COVID-19 patients. b, Quantification of lymphocytes using gating strategy shown in **Extended Data Fig. 1c** showed decreased amounts of T-cells in COVID-19 patients. c,d, Quantification of monocytes showed overall higher counts in COVID-19 patients that was due to higher number of classical monocytes (CD14^++^,CD16^−^), whereas non-classical monocytes(CD14^+^, CD16^++^) were reduced in COVID-19 patients. e-g, Analysis of marker expression on monocytes revealed reduced expression of HLA-DR (e), reduced number of CX3CR1 expressing monocytes (f) and increased number of CD11b expressing monocytes (g) in COVID-19 patients. h,i, Isolated PBMCs were stimulated with LPS (h) or R848 (i) for 24 hours after which production of IL-1β, IL-6 and TNFα was quantified in the supernatant using ELISA. COVID-19 patient PBMCs show increased cytokine production upon stimulation with either stimulus (n = 13 for COVID-19 patients, n = 10 for healthy controls). j, Isolated PBMCs were stimulated with heat-killed *Staphylococcus aureus* (HKSA) for 7 days after which production of IFNγ and IL-17 was quantified using ELISA. IFNγ response was reduced, whereas IL-17 production was elevated in COVID-19 patients. (n = 12 for COVID-19 patients, n = 10 for healthy controls) Data are presented as mean ± SEM.*p< 0.05, **p< 0.01, ***p< 0.001 for two-sided student’s ttest (for normally distributed data) or Kruskal-Wallis test.

**Fig. 2.**
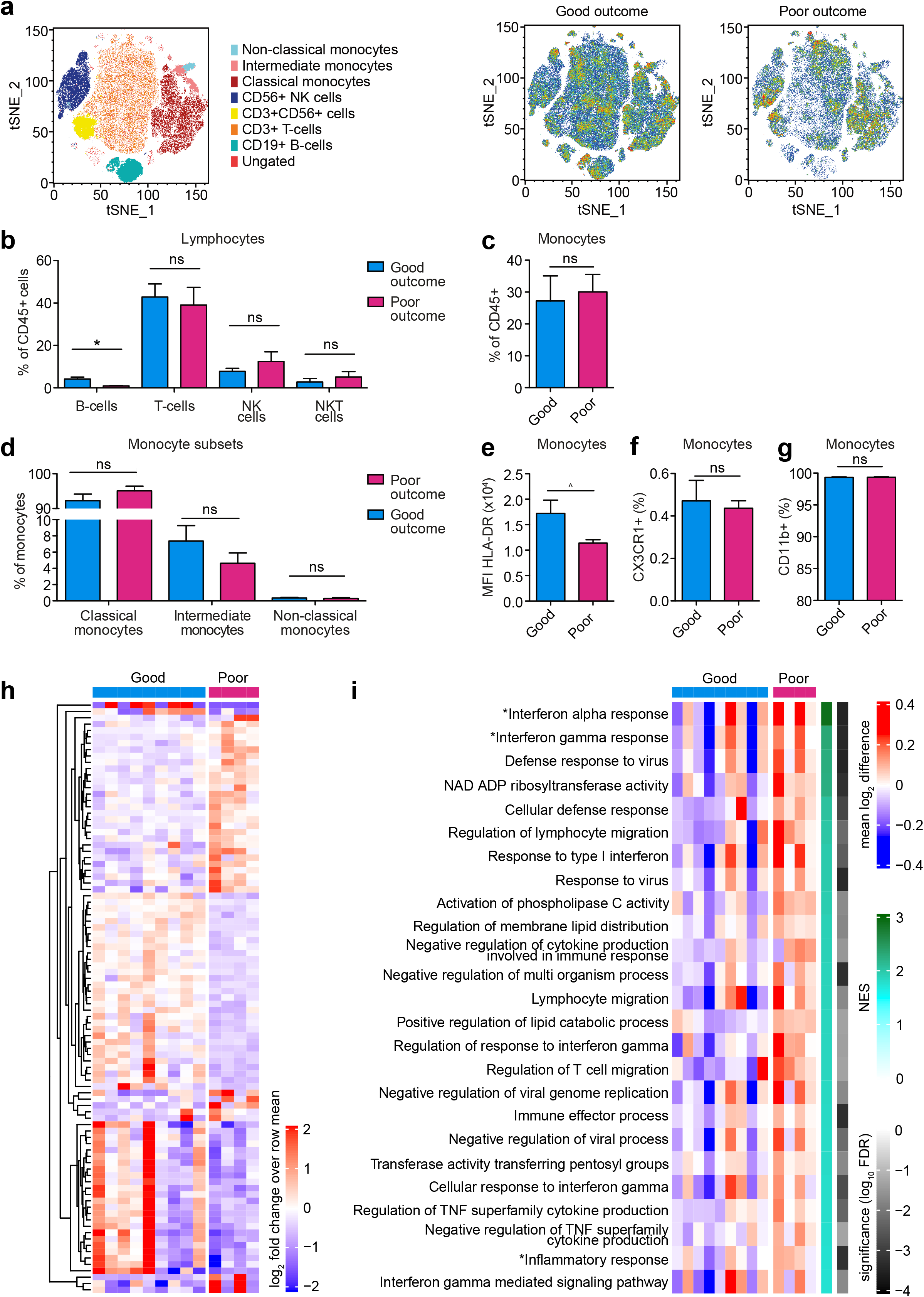
Innate immune response in COVID-19 patients at admission differs between patients with good or poor outcome. a-g, PBMCs isolated from COVID-19 patients at admission were analyzed using flow cytometry (n = 7 for COVID-19 patients with good outcome, n = 3 for COVID-19 patients with poor outcome). a, tSNE plots showing unsupervised clustering on the expression of 10 markers (CD45, CD14, CD16, CD3, CD19, CD56, HLA-DR, CD11b, CCR2 and CX3CR1) in COVID-19 patients with good and poor outcome. b, Quantification of lymphocytes using gating strategy shown in **Extended Data Fig. 1c** showed minor differences in B-cells between COVID-19 patients with good or poor outcome. c,d, Quantification of monocytes showed no difference between COVID-19 patients with good or poor outcome. e-g, Expression of HLA-DR (e), CX3CR1 (f) and CD11b (g) on monocytes did not differ between COVID-19 patients with good or poor outcome. h,i, Transcriptome analysis was performed on isolated monocytes of COVID-19 patients at admission. h, Heatmap of differentially expressed genes (p< 0.005) between COVID-19 patients with good and poor outcome. i, Heatmap of GO-pathways and hallmark-pathways that are significantly enriched (FDR< 0.05) in COVID-19 patients with good and poor outcome. (* HALLMARK pathways; NES: normalized enrichment score) Data are presented as mean ± SEM. ^p< 0.06, *p< 0.05 for two-sided student’s t-test (for normally distributed data) or Kruskal-Wallis test.

We performed functional assays by stimulating PBMCs for 24 hours *ex vivo* with subsequently measuring cytokine release, namely IL-1β, IL-6 and Tumor Necrosis Factor alpha (TNFα). We observed markedly elevated cytokine responses in COVID-19 patients upon Toll-like receptor (TLR) 4 activation by lipopolysaccharide (LPS) and TLR7/8 activation by R848 (**Fig. 1h,i**). Enhanced cytokine responses were also observed upon stimulation with the TLR2 agonist Pam3CSK4 and heat-killed *Candida albicans* (HKCA) (**Extended Data Fig. 1d,e**). This increased cytokine response was unchanged in patients who remained hospitalized throughout our five-day observation period (**Extended Data Fig. 2h**).

Next, we explored if the changes in the innate immune profile were associated with altered responses in the adaptive immune system. For this purpose, we stimulated PBMCs for 7 days with heat-killed *Staphylococcus aureus* (HKSA) and measured interferon-γ (IFNγ) as a marker of T helper cell (Th) 1 activation, and IL-17 as a marker of Th17 activation (**Fig. 1j**). In healthy controls we observed substantial IFNγ and little IL-17 production, indicating a dominant Th1 response. In contrast, IFNγ production was reduced and IL-17 production enhanced in COVID-19 patients, indicating polarization towards a Th17 response.

### Interferon-stimulated gene expression is related to development of severe disease

Of the 13 patients with COVID-19 that were included in our study, nine recovered without requiring ICU admission, while four had a poor outcome (ICU admission (n = 3) or death (n = 1)) (**Extended Data Fig. 1a**). At the time of presentation, we observed no clear differences in clinical variables between patients who would eventually have favorable outcomes and those who eventually had poor outcomes (**Extended Data Table 1**). We were interested if we could detect immune response differences at admission that could be related to patient outcome. No differences in leucocyte subsets between both groups was observed, except for a lower B-cell count in patients with poor outcome (**Extended Data Table 2; Fig. 2a-d**). Monocyte HLA-DR expression was reduced in patients with poor outcomes, indicating that an inflammatory monocyte phenotype was more pronounced in these patients (**Fig. 2e**). CX3CR1 and CD11b expression were equal in both groups (**Fig. 2f,g**). Next, we isolated monocytes of the COVID-19 patients and analyzed their transcriptomes by RNA sequencing. We found marked differences in transcription of interferon-stimulated genes, which are critical in the context of viral infections^16^. Notably, higher expression of interferon-stimulated genes was associated with poor outcome (**Fig. 2h,i, Extended Data Table 3**).

Six patients recovered fast and were discharged within the first five days, while seven patients remained hospitalized. We obtained PBMCs from this latter group five days after admission. At this timepoint, a clear distinction could be made, based on clinical parameters, between patients with good versus poor outcomes (**Extended Data Table 1**). No differences in lymphocyte subsets were observed (**Fig. 3a,b, Extended Data Fig. 3**). However, the poor outcome group had more classical monocytes (**Fig. 3c,d**). Importantly, the number of non-classical monocytes recovered in patients with good outcomes but remained virtually undetectable in deteriorating patients (**Fig. 3d**). Clear differences were visible in monocyte surface marker expression, with decreased HLA-DR and CX3CR1 expression in patients with poor outcomes (**Fig. 3e,f**). There was no difference in CD11b expression between the groups (**Fig. 3g**). Transcriptome analysis of circulating monocytes showed a clear distinction between patients with a good outcome and patients with a poor outcome. Similar to what we observed at the time of admission, we found enhanced transcription of interferon-stimulated genes five days after admission to be associated with poor outcome (**Fig. 3i,j**)

**Fig. 3.**
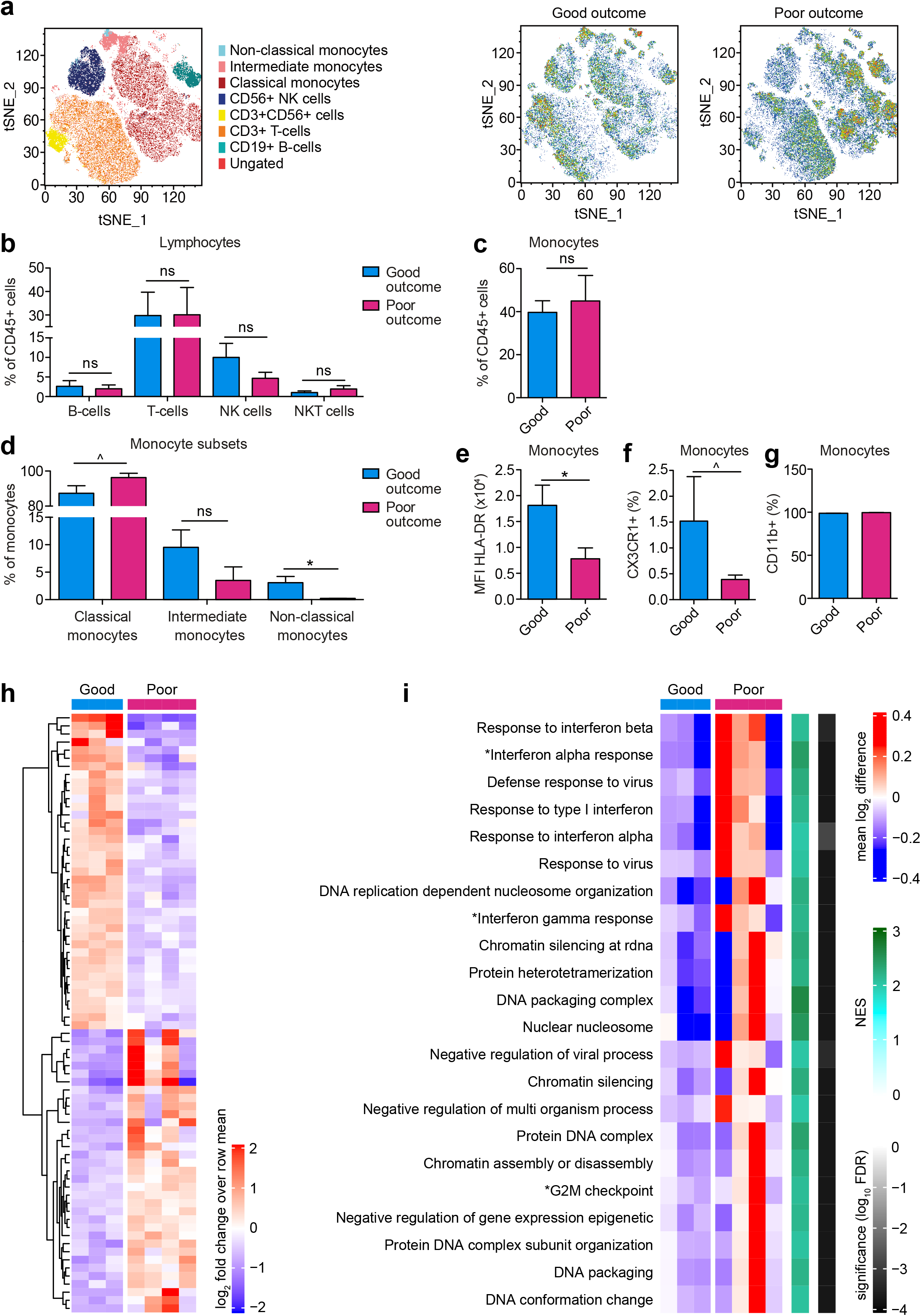
Differences in innate immune response in COVID-19 patients with good or poor outcome increases five days after admission. a-g, PBMCs isolated from COVID-19 patients five days after admission were analyzed using flow cytometry (n = 2 for COVID-19 patients with good outcome, n = 3 for COVID-19 patients with poor outcome). a, tSNE plots showing unsupervised clustering on the expression of 10 markers (CD45, CD14, CD16, CD3, CD19, CD56, HLA-DR, CD11b, CCR2 and CX3CR1) in COVID-19 patients with good and poor outcome. b, Quantification of lymphocytes using gating strategy shown in **Extended Data Fig. 1c** showed no differences between COVID-19 patients with good or poor outcome. c, Quantification of overall monocytes showed no difference between COVID-19 patients with good or poor outcome. d, Subset analysis of monocytes revealed increased amounts of non-classical monocytes (CD14^+^, CD16^++^) in COVID-19 patients with good outcome. e,f, Expression of HLA-DR (e) and numbers of CX3CR1 expressing monocytes (f) were increased in COVID-19 patients with good outcome. g, CD11b expression on monocytes did not differ between COVID-19 patients with good or poor outcome. h,i, Transcriptome analysis was performed on isolated monocytes of COVID-19 patients five days after admission. (n = 3 for COVID-19 patients with good outcome, n = 4 for COVID-19 patients with poor outcome) h, Heatmap of differentially expressed genes (p< 0.001) between COVID-19 patients with good and poor outcome. i, Heatmap of GO-pathways and hallmark-pathways that are significantly enriched (FDR< 0.01) in COVID-19 patients with good and poor outcome.(* HALLMARK pathways; NES: normalized enrichment score) Data are presented as mean ± SEM. ^p< 0.06, *p< 0.05 for two-sided student’s t-test (for normally distributed data) or Kruskal-Wallis test.

Taken together, these immune profiling data show that the inflammatory response in SARS-CoV-2 infection is characterized by marked alterations in the innate immune system, a result that corroborates previous reports^14^. Monocytes show signs of enhanced activation, and increased expression of interferon-stimulated genes, which are likely markers of disease severity as we found this to be associated with a poor outcome. Importantly, we revealed an elevated monocyte-derived cytokine response to *ex vivo* stimulation of TLR2, TLR4 TLR7/8. This enhanced responsiveness, which we observed to persist during the course of the disease, is reminiscent of the inflammatory phenotype previously reported in sepsis and influenza. While inflammation is beneficial if induced early in the infection, as it contributes to improved anti-viral mechanisms and elimination of infection, if exacerbated late during the course of the disease it may play a role in the development of the severe complications of COVID-19.

### Hydroxychloroquine prevents the induction of trained immunity

The data presented thus far revealed enhanced responsiveness of monocytes in patients with active COVID-19. Such functional adaptation of monocytes is also observed in processes like priming and trained immunity, which potentiate the anti-viral innate immune response^13,17^. This result prompted us to investigate whether 4-aminoquinolines can affect trained immunity. Chloroquine and hydroxychloroquine are weak bases that passively diffuse to the lysosome, where they interfere with its function^10^. Lysosomes are at the center of coordinating immunometabolism and the innate immune response via mammalian target of rapamycin (mTOR), which is activated at the lysosomal membrane (**Fig. 4a**)^18,19^. Interestingly, activation of key regulators of lysosome genes is characteristic of the trained macrophage phenotype and distinguishes it from its LPS-tolerized counterpart^20^.

**Fig. 4.**
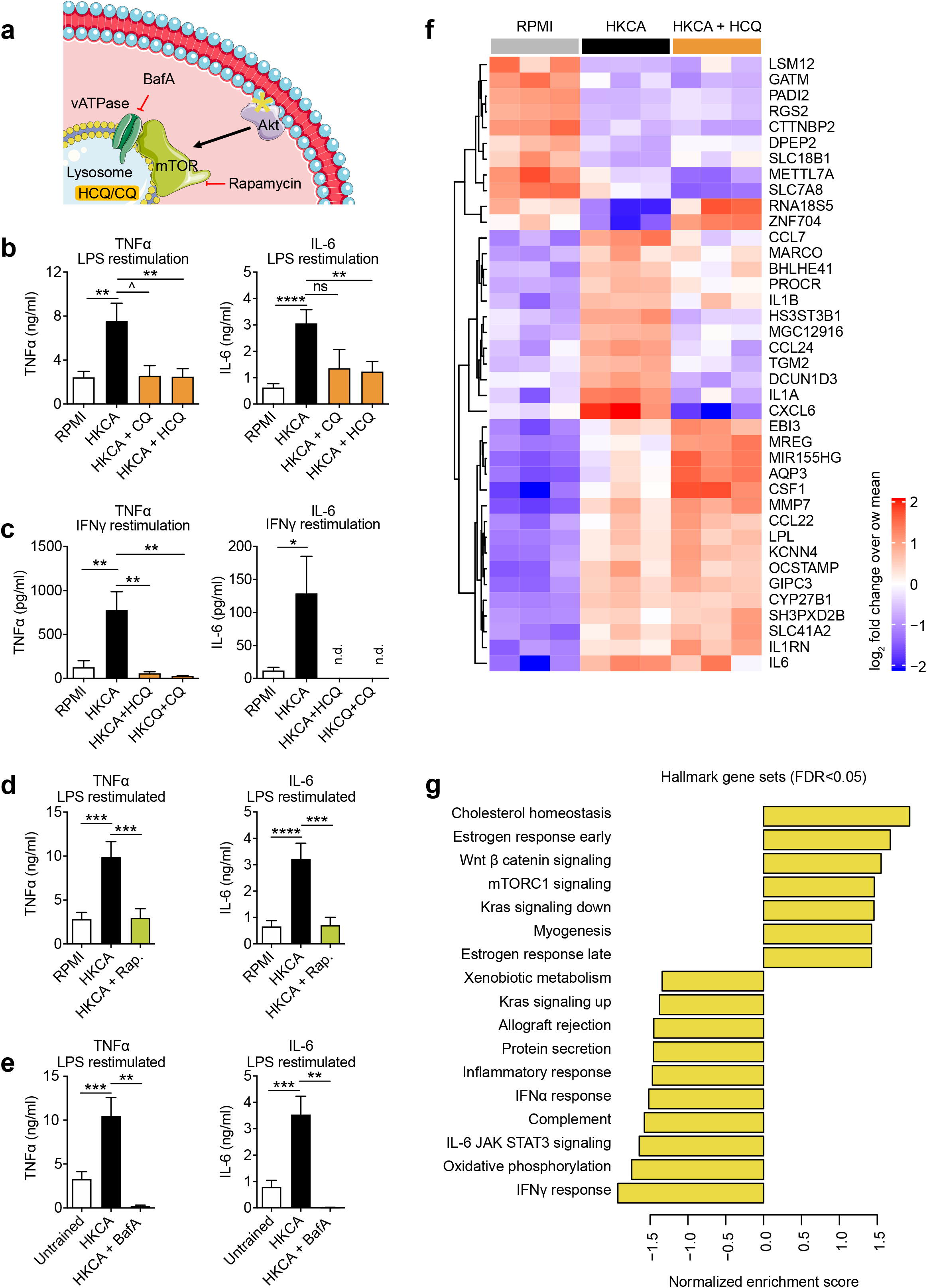
Hydroxychloroquine prevents the induction of trained immunity. a, Schematic representation of the interaction between the lysosome and the AKT/mTOR signaling pathway. b-e, PBMCs were stimulated for 24 hours with HKCA with or without specified inhibitors or RPMI as control. After a five-day resting period, cells were restimulated with LPS or IFNγ for 24 hours and cytokine production was measured in the supernatant. b,c, Hydroxychloroquine and chloroquine prevent induction of trained immune response to LPS (b, n = 7–17) and IFNγ (c, n = 5–8, n.d.:not detectable). d, mTOR inhibition prevents induction of trained immunity response to LPS (n = 11). e, Inhibiting lysosome acidification with Bafilomycin A prevents induction of trained immune response to LPS (n = 4–9). f,g, PBMCs were stimulated with HKCA, HKCA+HCQ or RPMI as control for 24 hours. Subsequently monocytes were purified and transcriptome analysis was performed. f, Heatmap of most significantly changing genes between HKCA stimulated and control PBMCs. g, Pathway analysis of most significant genes identified (FDR < 0.05) between HKCA treated cells and HKCA+HCQ treated cells. Normalized enrichment score is shown for HKCA+HCQ / HKCA, with positive values showing pathways upregulated in HKCA and negative values indicating pathways upregulated in HKCA+HCQ. Data are presented as mean ± SEM; ^p< 0.06, *p< 0.05, **p< 0.01, ***p< 0.001; One-way ANOVA with Dunnett’s post-test.

To investigate the effect of chloroquine (CQ) and hydroxychloroquine (HCQ) on trained immunity, we adapted a previously described *in vitro* protocol in which human PBMCs are stimulated with RPMI (control) or HKCA for 24 hours^21^. HKCA is a well described stimulus to induce trained immunity, but can also be induced by other stimuli such as IL-1β. The cells were subsequently washed and rested for five days in culture medium followed by a second 24-hour stimulus (LPS, Pam3CSK4 or IFNγ) (**Extended Data Fig. 4a**). We observed that HKCA-trained cells produced markedly more cytokines upon restimulation with either LPS or Pam3CSK4. This effect was abrogated when cells were treated with chloroquine and hydroxychloroquine for 24 hours during HKCA stimulation, indicating that these compounds prevent the induction of trained immunity (**Fig. 4b, Extended Data Fig. 4b**). Since interferons play a central role in viral immune responses, and our monocyte transcriptome data from COVID-19 patients revealed enhanced interferon-stimulated gene expression, we investigated how inflammatory monocytes respond to restimulation with IFNγ. Interestingly, we observed enhanced production of IL-6 and TNFα (**Extended Data Fig. 4c**). This effect was mitigated by chloroquine and hydroxychloroquine treatment during the HKCA training stimulus (**Fig. 4c**). We sought to assess if this was mediated through altered lysosomal function. Lysosomal proteins function in an acidic environment with a pH of around 4.5 to 5.0, which is maintained by the vacuolartype H+ –ATPase (V-ATPase). V-ATPase activity is also required for mTOR activation. Pharmacologic blocking of V-ATPase with Bafilomycin A1 prevented trained immunity, mirroring the effects of chloroquine and hydroxychloroquine as well as that of mTOR inhibition (**Fig. 4d,e, Extended Data Fig. 2d,e**).

Next, we investigated the transcriptomic effects of hydroxychloroquine treatment on trained monocytes. PBMCs were stimulated for 24 hours with either RPMI, HKCA or HKCA and hydroxychloroquine, after which we purified monocytes and performed RNA sequencing. hydroxychloroquine treatment significantly altered the monocyte transcriptome. Interestingly, hydroxychloroquine prevented the enhanced expression of genes that encode for IL-1α and IL-1β, which play central roles in trained immunity (**Fig. 4f**). Pathway analysis of differentially expressed genes revealed that hydroxychloroquine treatment substantially downregulated genes, including interferon-stimulated genes, related to inflammatory responses (**Fig. 4g**). We also observed distinct RNA expression patterns in metabolic pathways important for inflammation, namely those related to oxidative phosphorylation and cholesterol homeostasis (**Fig. 4g**). Altogether, these data indicate that hydroxychloroquine prevents the induction of trained immunity and suppresses expression of interferon-stimulated genes.

### Hydroxychloroquine affects the cellular lipidome

Our transcriptome data indicate that genes related to lipid metabolism play an important role in how hydroxychloroquine treatment prevents trained immunity. This corroborates previous studies in which the cholesterol synthesis pathway was shown to be involved in trained immunity^20,22^. We were interested in the monocyte lipidome in the context of trained immunity and the effect of hydroxychloroquine on this process. Accordingly, we analyzed the monocyte lipidome after 24 hours of HKCA stimulation with or without hydroxychloroquine by performing quantitative shotgun lipidomics, an unbiased mass spectrometry-based method that can detect hundreds of lipid types present in cells^23^. Principal-component analysis of the lipidomic data showed marked differences in the lipidomes of the HKCA-trained monocytes compared to trained monocytes treated with hydroxychloroquine and control monocytes (**Extended Data Fig. 5a**). Training with HKCA affected the concentrations of phosphatidylcholines (PC) and phosphatidylserines (PS) compared to control cells. Compared to HKCA training, hydroxychloroquine treatment altered a wide range of lipid classes, namely diacylglycerols (DAG), hexosylceramides (HexCer), alkyl-ether-linked lyso-phosphatidylcholines (LPC O-), lyso-phosphatidylethanolamines (LPE), alkyl-ether-linked lyso-phosphatidylethanolamines (LPE O-), phosphatidylcholines (PC), phosphatidylethanolamines (PE), phosphatidylglycerols (PG), phosphatidylinositols (PI), phosphatidylserines (PS) and triacylglycerols (TAG) (**Fig. 5a**). Interestingly, we observed that effects of HKCA training on various subspecies of PC, PE, PI and PS could be prevented by treatment with hydroxychloroquine (**Extended Data Fig. 5b-e**). Concentrations of PE and PC, which both can be synthetized from DAG, were reduced in HKCA-trained cells and remained at the level of control cells with hydroxychloroquine treatment (**Fig. 5f**). PS, which can be synthesized from PE and PC, was increased upon HKCA-training and remained at the level of control values with hydroxychloroquine treatment (**Fig. 5f**). A similar pattern could be observed for PI (**Fig. 5f**). In addition to the quantitative changes, lipid configurations were altered (**Fig. 5b-e**). Lipids isolated from HKCA-trained cells had longer acyl chains compared to lipids from hydroxychloroquine-treated and control cells (**Fig. 5b,c**) whereas no change in acyl chain length could be observed (**Fig. 5d**). HKCA-trained cells treated with hydroxychloroquine contained more lipids with saturated acyl chains (none, one or two double bonds) compared to HKCA-trained cells (**Fig. 5e**).

**Fig. 5.**
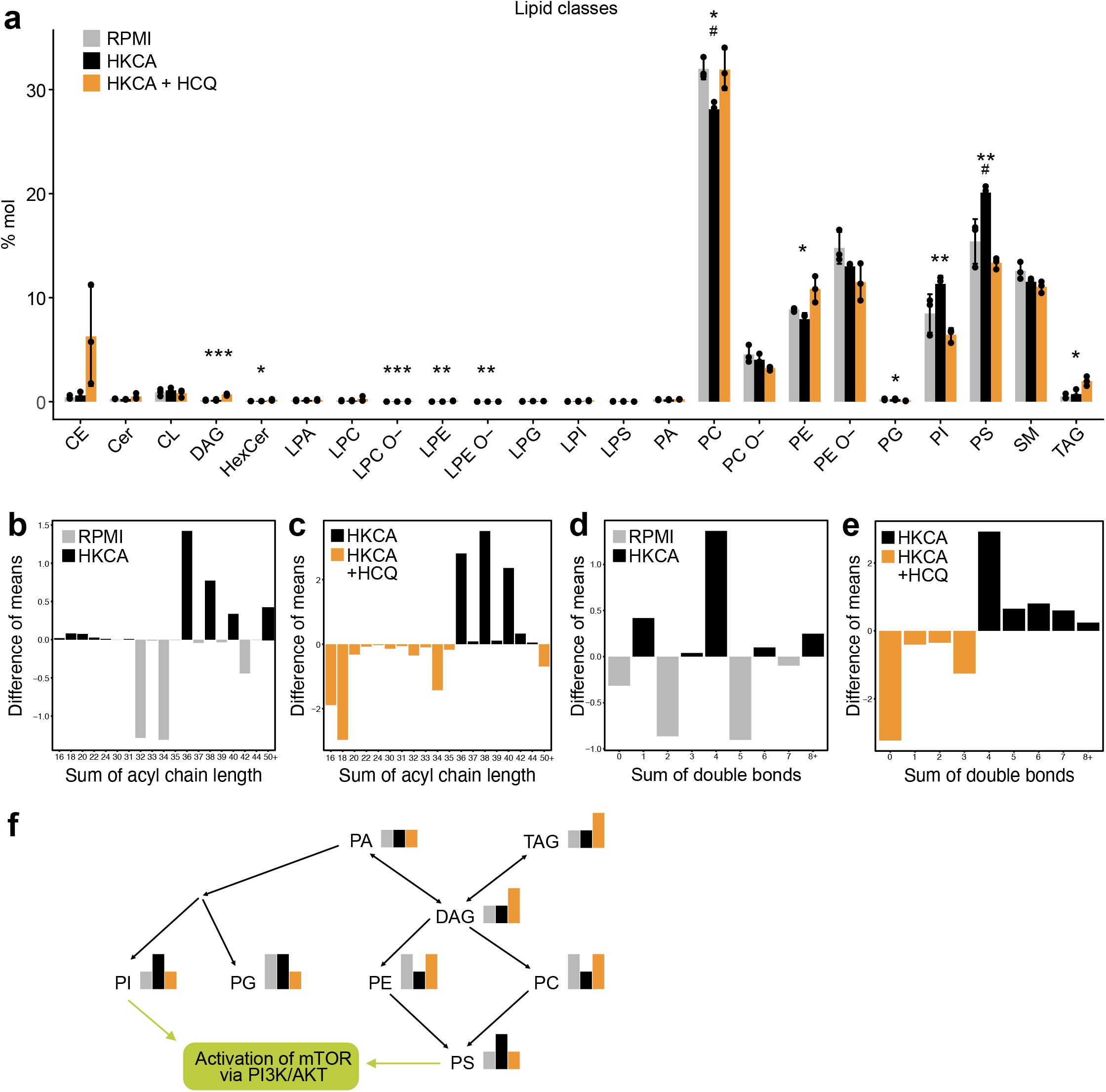
Hydroxychloroquine prevents the effect of heat-killed *Candida albicans*-training on the monocyte lipidome. a-e, PBMCs were stimulated with HKCA, HKCA+HCQ or RPMI as control for 24 hours. Subsequently monocytes were purified and analyzed for their lipid content using mass spectrometry-based shotgun lipidomics. (n = 3 per treatment group) a, Abundance of lipid classes as molar percentage of all lipids per treatment group. HKCA alone induced a significant decrease in phosphatidylcholines and an increase in phosphatidylserines compared to control cells, whereas HCQ induced significant changes in multiple lipid classes compared to HKCA treated cells. b,c, Analysis of acyl chain length of all lipids identified. HKCA-training resulted in lipids with longer acyl chains compared to control. HCQ induced even shorter acyl chains than control. d,e, Analysis of double bonds in all lipids identified. HKCA-training did not cause marked effects compared to control cells. HCQ-treated cells contained more lipids with fewer double bonds compared to HKCA-trained cells. f, Schematic representation of lipid metabolism showing lipid classes that are affected by either HKCA or HCQ treatment. Data are presented as mean ± SEM;^#^p< 0.05,^##^p< 0.01,^###^p< 0.001 between Control and HKCA; p< 0.05, **p< 0.01, ***p< 0.001 between HKCA and HKCA+HCQ; One-way ANOVA with Tukey post-test.

Our data indicate that trained immunity is accompanied by profound changes in the lipidome of monocytes. These changes may affect both cell and organelle membranes, thereby attenuating the function and activation of membrane-bound proteins. In this respect, it is interesting to note that PS and PI are essential to the activation of the phosphoinositide 3-kinase (PI3K)/AKT kinase complex, an important activation step in the mTOR pathway, and that mTOR itself requires the lysosomal membrane for its activation (**Fig. 4a**)^24–26^.

### Hydroxychloroquine prevents the epigenetic modifications necessary to induce trained immunity

Epigenetic changes provide the molecular substrate of trained immunity in monocytes and macrophages. As our functional assays showed that hydroxychloroquine prevents trained immunity, we investigated the impact of hydroxychloroquine on epigenetic regulation in monocytes. For this purpose, we performed a whole-genome assessment of the histone marks histone 3 lysine 27 acetylation (H3K27ac) and histone 3 lysine 4 trimethylation (H3K4me3) by chromatin-immunoprecipitation (ChIP) sequencing in control monocytes as well as HKCA-trained monocytes treated with or without hydroxychloroquine. Monocytes were trained as described previously, and after five days of rest monocyte-derived-macrophages were harvested for ChIP-sequencing. Epigenetic analysis by ChIP-seq of H3K27ac and H3K4me3 revealed marked differences between control and trained macrophages. After training and five days of resting, we found 352 peaks that had retained a significant change in H3K27ac and H3K4me3 dynamic between HKCA and control cells, indicating that an epigenetic memory was established. Interestingly, all HKCA-induced changes could be prevented with hydroxychloroquine treatment (**Fig. 6a**). Pathway analysis of differentially regulated peaks that remained active in HKCA-trained cells and were shut down in HKCA- and hydroxychloroquine-treated cells, revealed pathways associated with immune responses and inflammation (**Fig. 6b)**. These data therefore confirm our functional assays and show that hydroxychloroquine treatment effectively prevents the epigenetic changes underlying HKCA induced training and that this especially involves regulation of inflammation-related genes.

**Fig. 6.**
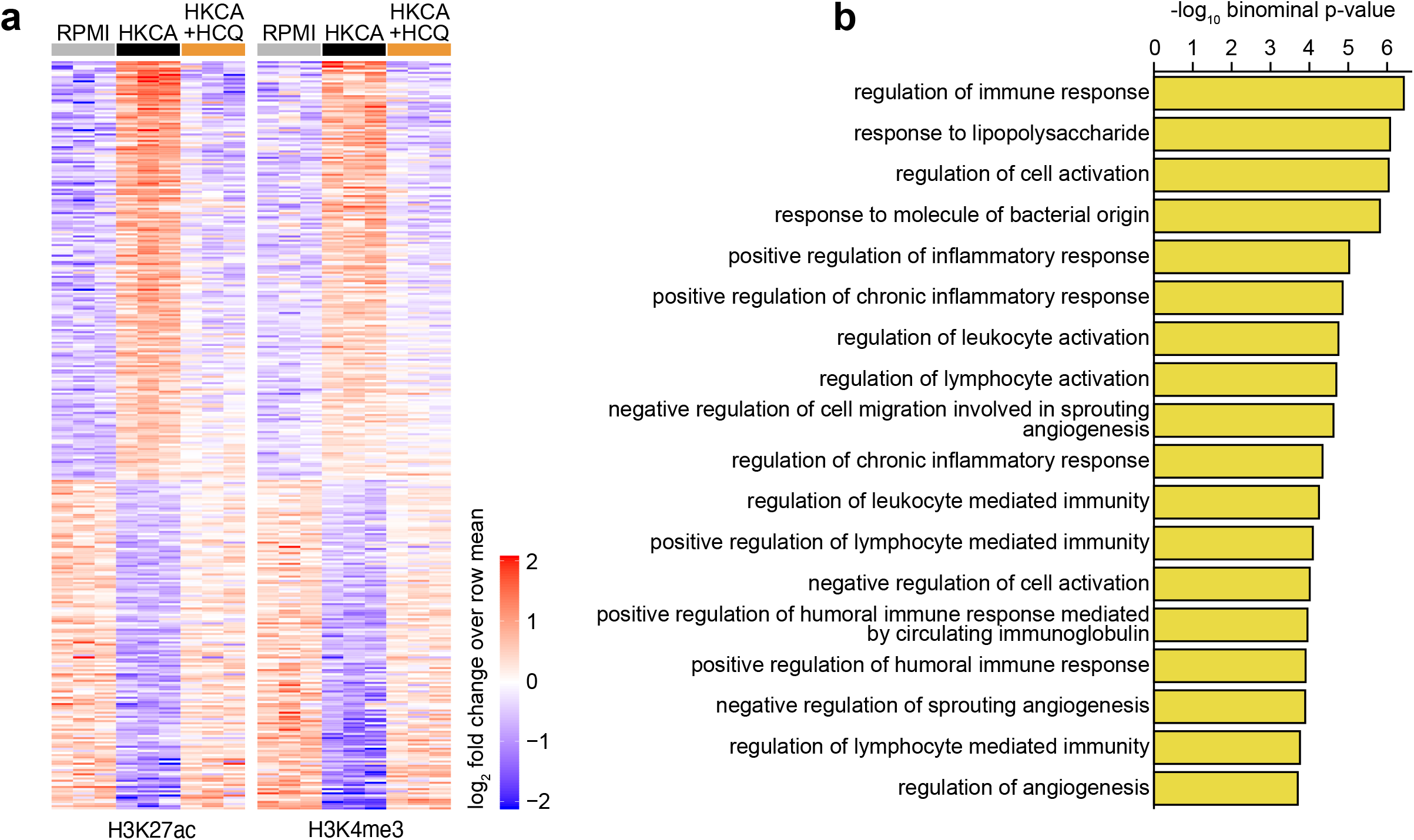
Epigenetic changes underlying heat-killed *Candida albicans* training are prevented by hydroxychloroquine. a,b, PBMCs were stimulated with HKCA, HKCA+HCQ or RPMI as control for 24 hours, after which cells were rested for five days. Subsequently monocytes were purified and chromatin-immunoprecipitation was performed for H3K27ac and H3K4me3. (n = 3 per treatment group) a, Heatmap showing relative H3K27ac (left panel) and H3K4me3 (middle panel) dynamics at sites with a significant change in histone mark abundance (FDR < 0.01) between HKCA stimulated and control monocytes. b, Gene ontology enrichment analysis of regulatory elements that remain epigenetically activated in HKCA-trained monocytes compared to HKCA+HCQ-treated cells (FDR < 0.01). Gene sets significantly associated at both an FDR Q value < 0.05 and a binominal p value < 0.05 are shown.

## DISCUSSION

SARS-CoV-2 infection primarily affects the upper respiratory tract and lung tissue. In most patients, an adequate immune response resolves the infection without causing organ damage. However, if the immune response is inadequate and viral clearance is impaired, severe pneumonia and other organ damage can develop, as is observed in patients with severe COVID-19^1^. Here we investigated the immune response in patients with COVID-19 and the immune-modulating action of hydroxychloroquine. We found that circulating monocytes from COVID-19 patients exhibit a phenotype of enhanced activation. Increased expression of interferon-stimulated genes by these cells associated with the development of more severe illness. We discovered that hydroxychloroquine can avert the induction of trained immunity at the functional and epigenetic level, possibly through changes in the cellular lipidome. As trained immunity comprises a functional adaptation of monocytes that enhances their anti-viral potential, this finding may have implications for the treatment of SARS-CoV-2 infection.

Our data provide a compelling new insight into the mechanism of action of hydroxychloroquine with important implications for its usefulness in COVID-19. Hydroxychloroquine’s immunomodulatory effects have been known for decades, and it is commonly used to prevent flares in rheumatic diseases, like systemic lupus erythematous and rheumatoid arthritis^10,27,28^. This drug can inhibit production of cytokines, like IL-1β, IL-6 and TNFα, by innate immune cells^29,30^. Yet the mechanism by which it inhibits cytokine production remains poorly understood. Hydroxychloroquine has a basic side chain and accumulates in the lysosome, where it exerts its effect, likely by impairing lysosome acidification rather than by targeting specific proteins. Previous studies support that hydroxychloroquine interferes with lysosomal processes, including autophagy^31^, antigen processing and major histocompatibility complex class II presentation^32,33^ as well as TLR7 and TLR9 processing and binding^34^.

We discovered a previously unknown immunomodulatory mechanism of hydroxychloroquine, namely that it prevents trained immunity through epigenetic modulation. This likely relates to lysosome acidification, as bafilomycin-A has a similar effect. We hypothesize that hydroxychloroquine may prevent trained immunity through effects on mTOR signaling, because mTOR closely interacts with the lysosome and is activated on its surface. Metabolic information from the lysosome is transmitted to the cell primarily through mTOR signaling^35^, which is key to mediating inflammation. A previous report demonstrated hydroxychloroquine’s effect on the mTOR pathway by showing that hydroxychloroquine decreased cellular levels of phospho-S6, a readout for mTOR activity^36^. Interestingly, hydroxychloroquine also has marked effects on lipid metabolism in monocytes. Changes in expression of genes involved in lipid metabolism were previously found to play an important role in trained immunity^20^. Our lipidomic studies showed that lipids belonging to the PI and PS class are upregulated upon HKCA training and that hydroxychloroquine treatment could prevent this increase. It is worth noting that mTOR activation via the Phosphoinositide 3-kinase (PI3K)/AKT pathway depends on the action of PS^26^, which brings AKT to the plasma membrane where it can be activated by PI’s^25^. Further studies are required to unravel the interaction between the lipidome and mTOR signaling in the context of trained immunity.

Our findings provide mechanistic insights that shed new light on the usefulness of hydroxychloroquine in COVID-19. We show that hydroxychloroquine prevents monocytes from adopting a trained immunity phenotype through effects on epigenetic reprogramming. Trained immunity is known to enhance the innate immune response and thereby facilitates the defense against infections. In fact, previous studies have shown that induction of trained immunity, e.g. through Bacillus Calmette-Guérin vaccination, can help prevent bacterial as well as viral infections^37,38^. The fact that hydroxychloroquine averts trained immunity argues against the usefulness of this drug in clearing SARS-CoV-2 infection. This corroborates the finding of a recent randomized controlled trial showing that hydroxychloroquine, when used as post-exposure prophylaxis, does not prevent symptomatic SARS-CoV-2 infection^12^. The question arises if the opposite, namely the induction of trained immunity, may actually be beneficial for this purpose. A randomized clinical trial is currently being conducted to investigate this^39^. A question that remains open is if hydroxychloroquine could be useful in severely ill COVID-19 patients. These patients are characterized by exaggerated inflammation which may contribute to disease pathology^2,3^, and hydroxychloroquine could theoretically reduce cytokine production. However, this drug may not be as potent as other anti-inflammatory drugs such as IL-6 receptor antibodies or the IL-1 receptor antagonist, which may be more effective treatment strategies. A recent observational study found no signs of benefit from hydroxychloroquine in the treatment of COVID-19^11^. Randomized clinical trials are needed to address this formally^40^.

In summary, we found that hydroxychloroquine averts the induction of trained immunity in monocytes through effects on the lipidome and epigenome. This occurred concomitantly with decreased expression of interferon-stimulated genes. Trained immunity comprises a functional adaptation that enhances the anti-viral potential of the innate immune system. Therefore, our findings suggest that hydroxychloroquine may not have a beneficial effect on the anti-viral immune response in SARS-CoV-2 infection.

## ONLINE METHODS

### Human Subjects

For *in vitro* studies on human PBMCs and monocytes, buffy coats from healthy donors were obtained from Sanquin blood bank, Nijmegen after written informed consent. Blood from COVID-19 patients was collected after written informed consent at Radboudumc. The study was approved by the local medical ethics committee of the Radboudumc under reference number: 2020–6359.

### Human PBMC isolation

PBMCs were isolated by differential centrifugation over Ficoll-Paque (Lymphoprep, Stemcell Technologies). Cells were washed three times in PBS. PBMCs and monocytes were resuspended in RPMI culture medium supplemented with 2mM glutamax, 1mM pyruvate and penicillin/streptomycin (all from Thermo Fisher Scientific) and counted on a Casy counter. Cell counts of whole blood and isolated PBMCs were also analyzed using a sysmex XN-450 automated hematology analyzer (Sysmex).

### Training and inhibition experiments

Human PBMCs were trained as described before. In short, 500.000 PBMCs were added into 96-well flat bottom plates. Cells were allowed to adhere for 1h at 37°C. Cells were washed three times with PBS prior to stimulations. After washing cells were incubated with culture medium only as negative control, or treated with 100 μM chloroquine (Sigma Aldrich), 100 μM hydroxychloroquine (Sigma Aldrich) or 0.01 μM rapamycin (Selckchem) for 1 hour at 37°C. Subsequently cells were incubated with 10^5^ cells/ml HKCA (Invivogen) for 24 hours together with the respective treatment for 24 hours at 37°C. Subsequently, cells were washed and cells were rested for five days in RPMI culture medium containing 10% FBS. After the resting period cells were stimulated with either RPMI as negative control, 10ng/ml LPS (Sigma Aldrich) or 1ug/ml Pam3CSK4 (Invivogen).

### PBMC stimulation of COVID-19 patients

PBMCs from COVID-19 patients were stimulated using 10 ng/ml LPS, 1 μg/ml Pam3CSK4, 10^6^ cells/ml HKCA and 10 μg/ml R848 (Invivogen) for 24 hours in RPMI without serum or with RPMI only as negative control or 10^6^ cells/ml heat-killed S*taphylococcus aureus* (ATCC) for 7 days in RPMI with 10% serum in 96-well round-bottom plates (Corning).

### Monocyte isolation

Monocytes were isolated using negative MACS isolation with the Pan monocyte isolation kit (Miltenyi Biotech). Briefly, stimulated PBMCs were washed with PBS and incubated with versene solution (0.48mM EDTA, Sigma Aldrich) for 30 minutes at 37 °C. Cells were scraped from the plates, counted, spun down and resuspended in MACS isolation buffer (PBS with 0.5% BSA and 2mM EDTA).

Monocytes from COVID-19 patients were isolated directly after isolation of PBMCs. PBMCs were counted, spun down and resuspended in MACS isolation buffer. Monocyte isolation was performed according to manufacturer’s instructions.

### Cytokine measurements

Cytokine production was measured in supernatants using commercial ELISA kits for human TNFα, IL-6, IFNγ, IL-22 and IL-17 (R&D systems) according to manufacturer’s instruction.

### Flow cytometry

Circulating immune cells and monocyte (sub)populations were identified by their expression markers using a CYTOflex flow cytometer (Beckman Coulter) (gating strategy in **Extended Data Fig. 1c**). Antibodies and dilutions used are CD45-BV510 (Biolegend, 1:100), CD14-PC7 (eBioscience, 1:100), CD16-FITC (eBioscience, 1:100), CD3-APC750 (Beckman Coulter, 1:50), CD19-APC-R700 (Becton Dickinson, 1:100), CD56-APC (Beckman Coulter, 1:50), HLADR-PE (Beckman Coulter, 1:20), CD11b-BV785 (Biolegend, 1:100), CCR2-BV421 (Becton Dickinson, 1:50), CX3CR1-BV650 (Biolegend, 1:50) and Live/Dead FVS620 (Becton Dickinson, according to manufacturers’ instructions). 500.000 PBMCs were stained with FVS620, subsequently underwent Fc blocking using 10% Heat-Inactivated human serum and were stained with antibodies in presence of Brilliant Stain buffer (Becton Dickinson) as multiple BV antibodies were used. Flow cytometry standards (FCS) files underwent pre-processing to remove debris, dead cells and doublets. Live single cells were then analyzed by both unsupervised computational analysis as well as manual gating in parallel. Characterization of monocytes subsets is according to current recommendations (See **Extended Data Fig. 1c**)^41^. For unsupervised computation analysis, FCS files were randomly down sampled to 20,000 events of the pre-processed files and subsequently concatenated to a single file containing all events. Controls and patients were labelled accordingly to be able to separate them after analysis. Unsupervised clustering was performed on the expression values of all markers using the tSNE plugin in FlowJo (Becton Dickinson, version 10.6.2), using 1000 iterations and a perplexity of 30. Manual gating of known cell populations (see gating strategy, **Extended Data Fig. 1c**) was used to identify populations and to check separation quality of the unsupervised clustering. The contribution of the control and patient populations to the total tSNE was then analyzed by separating the groups. Visual differences were then confirmed by manual gating and statistical analysis.

### RNA isolation, library preparation and sequencing for transcriptomic analysis

For RNA isolation 1*10^6^ isolated monocytes were resuspended in 350 μl of RNA later Buffer (Qiagen). RNA was isolated using RNeasy kit (Qiagen) including DNAseI (Qiagen) digestions. Total RNA isolated from monocytes was used for the preparation of the RNA sequencing libraries using the KAPA RNA HyperPrep Kit with RiboErase (KAPA Biosystems). In short, oligo hybridization and rRNA depletion, rRNA depletion cleanup, DNase digestion, DNase digestion cleanup, and RNA elution were performed according to protocol. Fragmentation and priming were performed at 94°C for 6 min. First strand synthesis, second strand synthesis and A-tailing was performed according to protocol. For the adapter ligation, a 1.5 μM stock was used (NextFlex DNA barcodes, Bioo Scientific). First and second post-ligation cleanup was performed according protocol. A total of 11 PCR cycles were performed for library amplification. The library amplification cleanup was done using a 0.8x followed by a 1.0x bead-based cleanup. Library size was determined using the High Sensitivity DNA bioanalyzer kit, and the library concentration was measured using the dsDNA High Sensitivity Assay (Denovix). Paired-end sequencing reads of 50 bp were generated using an Illumina NextSeq 500.

### Preparation of samples and lipid extraction for mass spectrometry lipidomics

For lipidomic analysis 10*10^6^ isolated monocytes were collected in to microcentrifuge tubes, centrifuged at 1000*x*g for 5 minutes at 4 °C. The supernatant was removed and cells were snap frozen in liquid nitrogen. Mass spectrometry-based lipid analysis was performed at Lipotype GmbH (Dresden, Germany) as described in^23^. Lipids were extracted using a two-step chloroform/methanol procedure^42^. Samples were spiked with internal lipid standard mixture containing: cardiolipin 16:1/15:0/15:0/15:0 (CL), ceramide 18:1;2/17:0 (Cer), diacylglycerol 17:0/17:0 (DAG), hexosylceramide 18:1;2/12:0 (HexCer), lysophosphatidate 17:0 (LPA), lyso-phosphatidylcholine 12:0 (LPC), lyso-phosphatidylethanolamine 17:1 (LPE), lyso-phosphatidylglycerol 17:1 (LPG), lyso-phosphatidylinositol 17:1 (LPI), lysophosphatidylserine 17:1 (LPS), phosphatidate 17:0/17:0 (PA), phosphatidylcholine 17:0/17:0 (PC), phosphatidylethanolamine 17:0/17:0 (PE), phosphatidylglycerol 17:0/17:0 (PG), phosphatidylinositol 16:0/16:0 (PI), phosphatidylserine 17:0/17:0 (PS), cholesterol ester 20:0 (CE), sphingomyelin 18:1;2/12:0;0 (SM) and triacylglycerol 17:0/17:0/17:0 (TAG). After extraction, the organic phase was transferred to an infusion plate and dried in a speed vacuum concentrator. 1st step dry extract was re-suspended in 7.5 mM ammonium acetate in chloroform/methanol/propanol (1:2:4, V:V:V) and 2nd step dry extract in 33% ethanol solution of methylamine in chloroform/methanol (0.003:5:1; V:V:V). All liquid handling steps were performed using Hamilton Robotics STARlet robotic platform with the Anti Droplet Control feature for organic solvents pipetting.

### Mass spectrometry data acquisition

Samples were analyzed by direct infusion on a QExactive mass spectrometer (Thermo Scientific) equipped with a TriVersa NanoMate ion source (Advion Biosciences). Samples were analyzed in both positive and negative ion modes with a resolution of Rm/z = 200 = 280000 for MS and Rm/z = 200 = 17500 for MSMS experiments, in a single acquisition. MSMS was triggered by an inclusion list encompassing corresponding MS mass ranges scanned in 1 Da increments^43^. Both MS and MSMS data were combined to monitor CE, DAG and TAG ions as ammonium adducts; PC, PC O-, as acetate adducts; and CL, PA, PE, PE O-, PG, PI and PS as deprotonated anions. MS only was used to monitor LPA, LPE, LPE O-, LPI and LPS as deprotonated anions; Cer, HexCer, SM, LPC and LPC O- as acetate adduct.

### Chromatin Immunoprecipitation

Isolated monocytes were resuspended in RPMI culture medium and fixed using formaldehyde (1% final concentration, Sigma Aldrich) for 10 minutes at room temperature. Unreacted formaldehyde was quenched with 125 mM glycine and incubated for 5 minutes at room temperature. Cells were washed twice in PBS containing protease inhibitor cocktail (Roche) and 1 mM PMSF (Roche), and subsequently snap frozen in liquid nitrogen. Cell pellets were stored at −80 °C for further use. Cells were sonicated at a concentration of 15 million cells/ml using a Bioruptor pico sonicator (Diagenode; 10 cycles, 30s on, 30s off, at 4 °C). Immunoprecipitation was performed using the MagnaChIP kit (Merck Millipore) according to manufacturer’s instruction. In short, 500,000 cells were incubated overnight with 1 μg H3K4me3 or H3K27Ac antibody (Diagenode) and protein A magnetic beads at 4 °C. Beads and chromatin/antibody mixture were washed four times for 5 minutes at 4 °C. After washing chromatin was eluted and proteins were degraded using proteinase K. DNA was purified using spin columns and eluted in millliQ.

### Library preparation and sequencing of ChIP samples

ChIP-seq libraries were prepared using the Kapa Hyper Prep Kit according to manufacturer’s protocol, with the following modifications. 2.5 μL of the NEXTflex adapter stock (600 nM, Bioo Scientific) was used for adapter ligation of each sample. Libraries were amplified with 12–15 PCR cycles followed by a double post-amplification clean-up was used to ensure proper removal of adapters. Samples were analyzed for purity using a High Sensitivity DNA Chip on a Bioanalyzer 2100 system (Agilent). Libraries were paired-end sequenced to a read length of 50 bp on an Illumina NextSeq500.

### *In vitro* experiments and flow cytometry data analysis

For *ex vivo* stimulations and flow cytometry data, data are shown as mean ± SEM and significance is tested using two-sided student’s t-test (for normally distributed data) or Kruskal Wallis. For *in vitro* trainings-experiments, data is shown as mean ± SEM and significance is tested with one-way ANOVA with Dunnett’s post-test. Data was analyzed using GraphPad Prism 5.0. P value of 0.05 was considered to be statistically significant.

### RNA-seq data analysis

RNA-sequencing reads were aligned with Hisat2 version 2.0.4 to the provided and pre-indexed hg38 transcript assembly from UCSC, with alignment tailoring for transcript assemblers enabled^44^. Samtools was used to filter reads with a quality score lower than 20, and PCR duplicates were removed with Picard^45^. Reads per gene were counted with the htseq-count script from the Hisat2 software suite using the GTF file corresponding to the transcript assembly, with reverse strandness enabled and identification attribute set to gene_id. Differential gene expression analysis was performed with the DESeq2 package version 1.18.1^46^. Genes with no reads mapped in any of the samples were filtered prior to differential expression analysis. The ‘rlogTransformation’ function in DESeq2 was used to normalize, transform and noise-stabilize the expression data for visualization purposes. All analyses were performed in R and figures were generated with the ggplot2 and ComplexHeatmap R packages^47–49^.

Gene set enrichment analysis was performed with the fgsea R package, by ranking genes with the ‘lfcShrink’ function in DESeq2^46,50^. The Hallmark and Gene Ontology (GO) gene set databases from MSigDB v6.2 were tested for significant associations with prognosis or response^50^. At least 1,000,000 permutations were performed to control for the false discovery rate, and the minimal and maximum size of gene sets to be considered for analysis were set to 15 and 500 genes, respectively. To prevent masking of potential heterogeneity within patient groups, gene set enrichment analysis results were visualized by plotting the mean gene expression change of all detected genes in a significant gene set for each sample separately.

### ChIP-seq data analysis

ChIP sequencing data was aligned to human genome hg19 with BWA^51^. Samtools was used to filter reads with a quality score lower than 20, and PCR duplicates were removed with Picard^45^. Peaks were identified with MACS 2.2.6 in paired-end mode and ‘call-summits’ enabled at a false discovery rate of 0.01^52^. A union of all identified peaks was generated with BEDTools, which was used to count reads per peak in each sample^53^. Read counts were analyzed with DESeq2 to identify significant dynamics, as described for the RNA-seq analysis. We used GREAT to identify significantly associated gene ontologies, and to assign each ChIP peak to its closest gene for integration between ChIP- and RNA-seq data^54^.

### Lipidomics data analysis

Data were analyzed with in-house developed lipid identification software based on LipidXplorer^55,56^. Data post-processing and normalization were performed using an in-house developed data management system. Only lipid identifications with a signal-to-noise ratio > 5, and a signal intensity 5-fold higher than in corresponding blank samples were considered for further data analysis. Further data analysis was performed using a web-based analysis program lipotypeZoom. Figures were generated in R with the ggplot2 and ComplexHeatmap R packages^47–49^. Data are shown as mean ± SEM, significance was determined using one-way ANOVA and Tukey post-test.

## Data Availability

Please contact corresponding author for data requests.

## ACKNOWLEDGEMENTS

This work was supported by Radboudumc Hypatia grant (to R.D.). MGN was supported by an ERC Advanced grant (833247) and a Spinoza grant of the Netherlands Organization for Scientific Research. SB is supported by the Dutch Heart Foundation (Dekker grant 2018-T028). The Vermeulen lab is part of the Oncode Institute, which is partly funded by the Dutch Cancer Society (KWF).

## AUTHOR CONTRIBUTIONS

N.R. and R.D. designed the study. N.R., C.Y., R.L., S.B., M.M.T.v.L., B.B., I.J., M.d.G., M.B., L.L., N.P.R., Z.A.F., W.J.M.M., L.B.H., L.A.B.J., M.G.N., M.V., J.v.d.V. and R.D. designed, performed and oversaw *in vitro* and *ex vivo* experiments. N.R., C.Y., B.B., M.d.G. and R.D. collected COVID-19 patient samples. I.J and R.D. collected and analyzed clinical characteristics of COIVD-19 patients. N.R. and R.D. collected and prepared samples for lipidomic analysis. M.B. and L.L. prepared samples for and performed RNA and CHIP sequencing. RNA and ChIP sequencing data analysis was performed by R.L and M.V. S.B. and C.Y. performed and analyzed flow cytometric study of COVID-19 PBMCs. The manuscript was written by N.R. and R.D. All authors contributed to writing of the manuscript and approved the final draft. R.D. provided funding.

## COMPETING INTEREST STATEMENT

The authors declare no competing interests.

## Notes

### Competing Interest Statement

The authors have declared no competing interest.

### Author Declarations

Blood from COVID-19 patients was collected after written informed consent at Radboudumc. The study was approved by the local medical ethics committee of the Radboudumc under reference number: 2020-6359.

